# Association of adverse childhood experiences with subsequent kidney disease among middle-aged and older adults in China: A national analysis

**DOI:** 10.1101/2022.06.08.22276145

**Authors:** Wenming Shi, Yonggang Huang, Changbo Jin

## Abstract

**Objective:** Few studies have been performed to address the impacts of adverse childhood experiences (ACEs) on kidney function in later life. We aimed to investigate the association between ACEs with subsequent kidney disease among middle-aged and older adults.

**Methods:** This national population-based study used data from the China Health and Retirement Longitudinal Study (CHARLS) 2015 and the life history survey in 2014. A total of 10102 participants aged ≥ 45 years from China were included. A wide range of 11 ACE indicators including childhood hunger, child’s poor health, physical abuse, emotional neglect, loneliness, peer bullying, domestic violence, household mental illness, household substance abuse, parental death, and incarcerated household member were measured by validated questionnaires. The cumulative number of ACEs was summed and and classified into four subgroups: ≤ 1, 2, 3 and ≥ 4. Glomerular filtration rate (eGFR) was estimated by serum cystatin C concentration and chronic kidney disease (CKD) was defined as eGFR < 60 ml/min/1.73m^2^. Multiple regression models were used to explore the relationship between accumulated ACEs and individual ACE indicator with subsequent kidney function.

**Findings:** Of the 10102 participants, 46.8% were males, and 16.0% reported exposure to four or more ACEs. Compared those with ≤ 1 ACE, participants who experienced four or more ACEs have a higher risk of decreased eGFR (β= -1.169, 95%CI: -2.113 to -0.225) and CKD (adjusted odds ratio, aOR=1.35, 1.04-1.75), after controlling for confounders. Exposure to specific ACE indicators of childhood poor health, physical abuse and household mental illness presented significant associations with reduced eGFR. The effects were more evident in men aged ≥ 60 years, with lower education or worse financial status.

**Conclusions:** Our study suggests that higher ACEs exposure increased the risk of subsequent kidney disease. The findings provide implications for mitigating the adverse effects of early-life stress and promoting kidney health by reducing ACEs.

## Introduction

Chronic kidney disease (CKD), one of the most challenging global health problems in the world, is an outcome of long-term accumulation of kidney injury(1). With the population aging, the prevalence of CKD is increasing worldwide and is linked to growing mortality, morbidity and healthcare expenditures(2). A national CKD survey in China showed that there were about 119.5 million patients with CKD during 2007-2010(3). Glomerular filtration rate (GFR) is widely used for kidney function assessment, however, the actual measurement of GFR is more difficult. Therefore, the GFR equation is often calculated to estimate the eGFR in clinical practice(4).

Studies have indicated besides the well-known risk factors for CKD, such as age, smoking, hypertension, and diabetes(5, 6), the early-life stressors including adverse childhood experiences (ACEs) may be a potential contributor to the risk of CKD (7-9). ACEs refer to a range of intensive stressors that occur before the age of 18 that have been shown to be associated with the risk of chronic diseases and early mortality(10, 11). According to the “developmental origins of health and disease” (DOHaD) theory, exposure to adverse events during a sensitive developmental period might lead to toxic stress response(9, 12, 13). This ensuing response was central to the development of chronic diseases, and enhancing the disease burden from a life-long perspective. Specific to kidney disease, several studies reported that the prevalence and risk of kidney disease was higher among the individuals who experienced high ACEs(9, 12, 13). An epidemiological study in China showed that exposure to childhood trauma such as hunger or food deprivation from famine was related with an increased risk of CKD in adulthood(8). Nevertheless, few studies have explored the long-term effects of different ACE indicators exposure on risk of kidney disease later in life. Additionally, little is known about the sociodemographic characteristics, i.e. gender, age and educational level, modify the association of early-life exposure to ACEs and kidney disease in elderly. To build upon existing literature, a large-scale study with clustering of ACE indicators to explore these relationship is timely needed.

In this study, we used a wide range of 11 ACE indicators from the China Health and Retirement Longitudinal Study (CHARLS), a national representative survey on middle-aged and older adults, to investigate the relationship between early-life exposure to multiple ACE indicators with the risk of kidney disease in later life. In addition, we also examined whether the association was modified by sociodemographic characteristics of age, gender, educational level and family’s financial status during childhood.

## Methods

### Study design and participants

This study used data from the China Health and Retirement Longitudinal Study (CHARLS), a nationally representative survey of 45 years and older adults from 28 provinces across mainland China. The details of the study design and sampling method have been described in previous literature(15). Briefly, a total of 17708 respondents were recruited by a multistage probability sampling strategy during the baseline survey, and follow-up every two years. Information on sociodemographic characteristics, behaviour and lifestyles, chronic conditions, and blood testing were collected by trained interviewers. In this study, we used the third wave of CHARLS (2015) project consisting of 21095 respondents, and then matched to the 2014 CHARLS life history survey (N= 20544). A total of 18678 individuals who completed both two surveys were included. After excluding individuals with missing data on blood testing, ACE indicators and other confounders, totally 10102 participants aged ≥ 45 years from China were enrolled in our final analysis. Ethics approval was obtained from the Institutional Review Board of Peking University. All participants provided their written informed consents. Figure S1 showed the flowchart of the participants’ enrollment.

### Kidney function measurement

Kidney function is usually assessed by eGFR through the CKD Epidemiology Collaboration (CKD-EPI) equation. However, several previous studies reported that gender, age, race, and protein intake can impact serum creatinine (Cr) concentration, it might result in a biased estimation of eGFR(15). In this study, we adopted to use the 2012 CKD-EPI cystatin C (CKD-EPICysC) equation (17). Compared to the CKD-EPI equation, the 2012 CKD-EPICysC equation is almost unaffected by aforementioned factors, and is suggested to be a potential alternative to creatinine for eGFR estimation. In the CHALRS project, all blood samples of participants were collected and immediately frozen at -20□ and transported to the China Center of Disease Control within two weeks where they were stored at -80□ until assay at local laboratory(18). Serum CysC concentration was tested by particle-enhanced turbimetric assay at the Youanmen Center for Clinical Laboratory of Capital Medical University. The coefficients of variation were controlled at 5% both within-assay and between-assay(18). CKD was defined by the eGFR less than 60 ml/min/1.73m^2^.

### Definition of adverse childhood experiences

We used a wide range of 11 ACE indicators from the CHARLS project, including childhood food deprivation, childhood poor health, physical abuse, emotional neglect, loneliness, peer bullying, domestic violence, household mental illness, household substance abuse, parental death, and incarcerated household member. Table S1 presented the details about the definition and measurement of each ACE indicator. Response to each indicator was dichtomized (0 vs.1) and then calculated to generate a cumulative score for each participant, ranging from 0 to 11. We classified the participants into four subgroups according to accumulated ACE score: ≤1 (used as reference), 2, 3 and ≥ 4.

### Covariates

Information on sociodemographic characteristics, chronic health conditions, and behaviour factors were collected by a validated CHARLS questionnaire. The sociodemographic characteristics included age, gender, body mass index (BMI), educational level, marriage status, and childhood financial status before 17 years (which evaluated by asking the participants “ When you were a child before the age of 17 years, compared to the average family in the same community/village, how was your family’s financial situation?” and was divided to two groups: same as or better than others vs. worse than others). The behaviour factors including cigarette smoking status (current; former: quitted smoking ≥ three years; never), and alcohol use (yes vs. no) were also recorded. The chronic conditions such as hypertension, diabetes and heart diseases were collected based on clinical and laboratory diagnosis or self-reported records. Hypertension was defined as systolic blood pressure (SBP) ≥ 140 mmHg and/ or diastolic BP (DBP) ≥ 90 mmHg or currently taking antihypertensive medicines (19). Diabetes was defined as fasting glucose ≥ 126 mg/dL, and/or glycosylated hemoglobin (HbA1c) ≥ 6.5%, and/or insulin use according to the recommendations of the American Diabetes Association (20).

### Statistical analyses

Descriptive statistic analysis were performed for all sociodemographic characteristics, behaviour factors and chronic conditions data. The continuous variables and classified variables were characterized by mean ± standard deviation (SD) and numbers with ratios, respectively. The prevalence of each ACE indicator and accumulated ACEs were calculated. Generalized linear regression and logistics regression models were performed to investigate the association between early-life exposure to accumulated ACEs and risk of the reduced kidney function or CKD, compared to the reference group (with ≤ 1 ACE). We fitted three models: model □ was a crude model; model □ adjusted for gender, age, BMI, marriage status, educational level, childhood financial status before 17 years, cigarette smoking status, and alcohol use; model □ further adjusted for hypertension, diabetes, and heart disease based on model □. The β-coefficient or odds ratio (OR) and 95% confidence intervals (95%CI) were calculated to report the association. We also tested the interaction of accumulated ACEs with demographic and economic covariables. Trend analyses were used to explore whether a dose-response relationship was present.

To identify the potential modifiers of several sociodemographic variables, stratified analyses were performed for gender (men vs. women), age (≥ 60 years vs. < 60 years), educational level (≥ middle school vs. < middle school) and family’s financial status before 17 years (same as or better than others vs. worse than others). We also evaluated the interaction of accumulated ACEs and each aforementioned variable in the regression models. The differences of effects between the paired strata were tested as following:

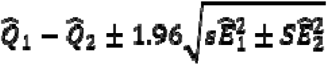

where 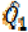 and 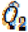 were the adjusted estimates for the two categories, and 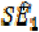 and 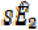 were the standard errors(21, 22).

All statistical analyses were conducted using STATA 16.0 (Stata Corp., College Station. TX, USA). All *p-*values were two sided, and significance was set as *p* < 0.05.

## Results

Of the 10102 participants included, 4731(46.8%) were men, and the average age was 61.22 ± 9.10 years. The sociodemographic characteristics, behaviour factors and chronic diseases according to the number of ACEs were summarized in Table 1. Overall, 67.9% of the participants reported experiencing two or more ACEs, and 16.0% had four or more ACEs (Table 1). Compared to those with ≤ 1 ACE, individuals who experienced four or more ACEs were more likely to be older, men, of lower educational level, of worse financial status during childhood, and more likely to have current smoking (Table 1). The average level of eGFR was lower in individuals with two or more ACEs groups than the reference group (P< 0.001, Table 1)

**Table 1.** The sociodemographic with behaviour characteristics and chronic conditions of participants by number of ACEs [Mean ±SD or N (%)].

| Variables | ACE score |  |  |  | Total<br>(N=10102) | P-value |
| --- | --- | --- | --- | --- | --- | --- |
|  | ≤ 1<br>(N=3242) | 2<br>(N=3056) | 3<br>(N=2189) | ≥ 4<br>(N=1615) |  |  |
| Age (years) | 60.49 ± 9.43 | 61.61 ± 9.11 | 61.43 ± 8.77 | 61.66 ± 8.78 | 61.22 ± 9.10 | <0.001 |
| Men (%) | 1445 (45.6) | 1376 (45.0) | 1071(48.9) | 839 (52.0) | 4731(46.8) | <0.001 |
| BMI (kg/m<sup>2</sup>) | 24.09 ± 3.78 | 23.91 ± 3.78 | 23.71 ± 3.66 | 23.58 ± 3.72 | 23.88 ± 3.75 | <0.001 |
| Educational level |  |  |  |  |  |  |
| ≥ Middle school | 1205 (37.2) | 889 (29.1) | 680 (31.1) | 466 (28.9) | 3240 (32.1) | <0.001 |
| < Middle school | 2037 (62.8) | 2167 (70.9) | 1509 (68.9) | 1149 (71.1) | 6862 (67.9) |  |
| Marriage status |  |  |  |  |  |  |
| Married | 2815 (86.8) | 2637 (86.3) | 1891(86.4) | 1389 (86.0) | 8732 (86.4) | 0.863 |
| Unmarried | 427 (13.2) | 419 (13.7) | 298 (13.6) | 226 (14.0) | 1370 (13.6) |  |
| Financial situation before 17 years |  |  |  |  |  |  |
| Same as or better than others | 2207 (68.1) | 1824 (59.7) | 1182 (54.0) | 787 (48.7) | 6000 (59.4) | <0.001 |
| Worse than others | 1035 (31.9) | 1232 (40.3) | 1007 (46.0) | 828 (51.3) | 4102 (40.6) |  |
| Cigarette smoking status |  |  |  |  |  |  |
| Current | 822 (25.4) | 829 (27.1) | 657 (30.0) | 481 (29.8) | 2789 (27.6) | <0.001 |
| Never | 1996 (61.6) | 1822 (59.6) | 1246 (56.9) | 881 (54.6) | 5945 (58.8) |  |
| Former | 424 (13.1) | 405 (13.3) | 286 (13.1) | 253 (15.7) | 1368 (13.6) |  |
| Alcohol use |  |  |  |  |  |  |
| Yes | 1146 (35.3) | 999 (32.7) | 791 (36.1) | 562 (34.8) | 3498 (34.6) | 0.045 |
| No | 2096 (64.7) | 2057 (67.3) | 1398 (63.9) | 1053 (65.2) | 6604 (65.4) |  |
| Chronic conditions |  |  |  |  |  |  |
| Hypertension (%) | 958 (29.5) | 939 (30.7) | 631 (28.8) | 464 (28.7) | 2992 (29.6) | 0.380 |
| Diabetes (%) | 592 (18.3) | 523 (17.1) | 375 (17.1) | 279 (17.3) | 1769 (17.5) | 0.599 |
| Heart disease (%) | 391 (12.1) | 392 (12.8) | 280 (12.8) | 216 (13.4) | 1279 (12.7) | 0.591 |
| Kidney biomarkers |  |  |  |  |  |  |
| Cysc (mg/l) | 0.84 ± 0.24 | 0.86 ± 0.27 | 0.86 ± 0.23 | 0.87 ± 0.25 | 0.86 ± 0.25 | 0.001 |
| eGFR (ml/min/1.73m<sup>2</sup>) | 93.19 ± 19.24 | 91.38 ± 19.19 | 91.83 ± 18.89 | 90.62 ± 19.89 | 91.94 ± 19.28 | <0.001 |

The prevalence of each specific ACE indicator ranged from 0.24% (incarcerated household members) to 70.9% (childhood food deprivation). Figure 2 presented the prevalence of each item of individual ACE indicators and accumulated ACEs. The multiple regression model showed that individuals who experienced four or more ACEs had a significantly higher risk of reduced eGFR in adults, when compared with those with ≤ 1 ACE. After controlling for the related confounders, the adjusted β and 95%CI were -1.169 (−2.113 to -0.225) for the decreased kidney function. Regarding CKD, we observed consistently significant association with aOR and 95%CI at 1.35 (1.04 to 1.75) (Table 2). The trend analysis showed a significant dose-response relationship between accumulative ACEs exposure and the decreased eGFR (P_trend_ = 0.023, Table 2). No statistical significance was observed for the interaction between accumulated ACEs with sociodemographic variables (P all > 0.05, data not shown). Specific to each individual ACE indicator, we found that participants who having poorer health during childhood (β= -1.196, 95%CI: -2.114 to -0.280) or with household mental illness (β= -0.916, 95%CI: -1.823 to -0.009) increased the risk of subsequent kidney disease (Figure 2). Furthermore, we also observed that early-life exposure to physical abuse was significantly related to an increased risk of kidney disease later in life. However, there was no significant association for other ACE indicators (Figure 2).

**Figure 1.**
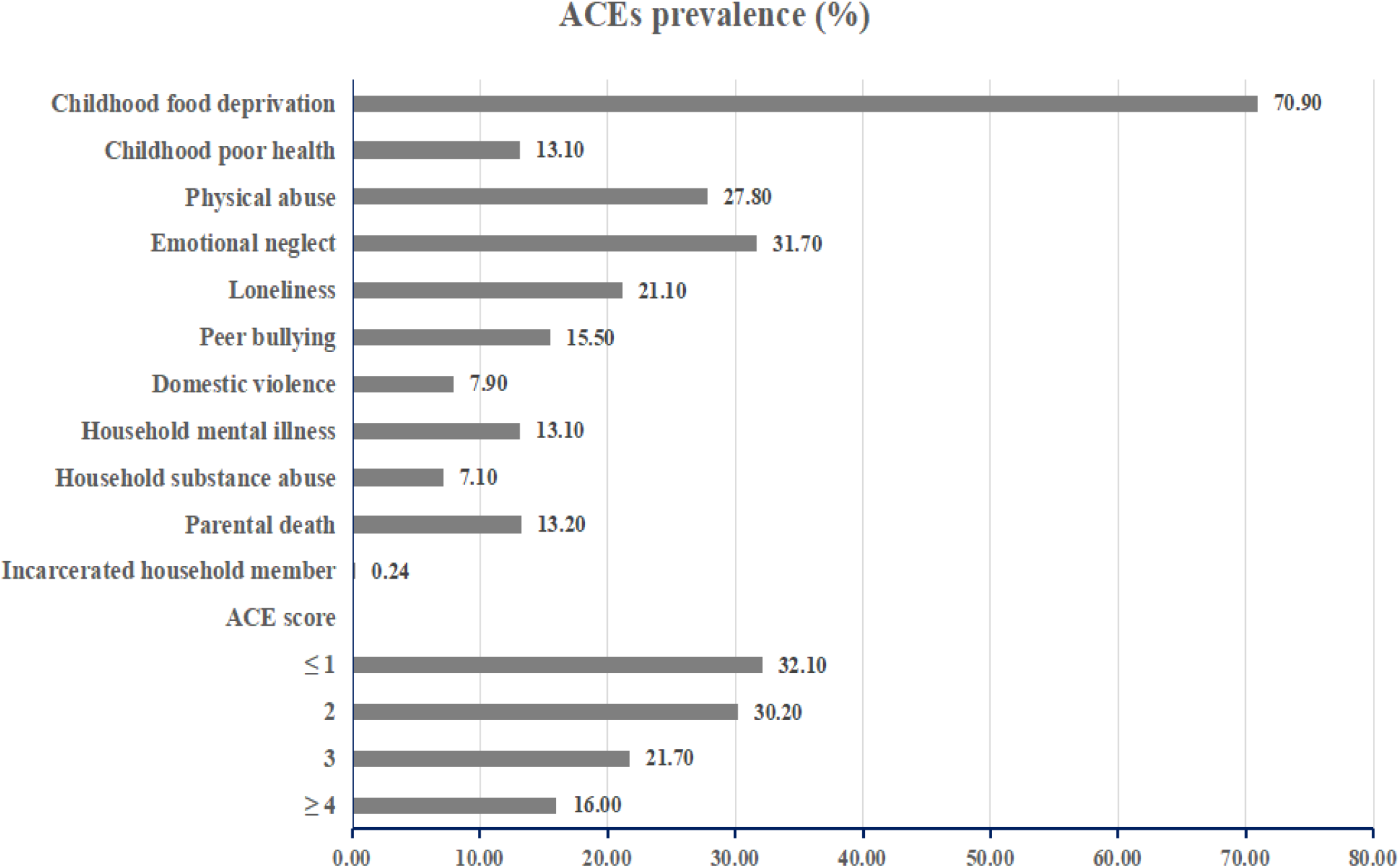
The prevalence of each ACE indicator and the accumulated ACEs among the participants.

**Figure 2.**
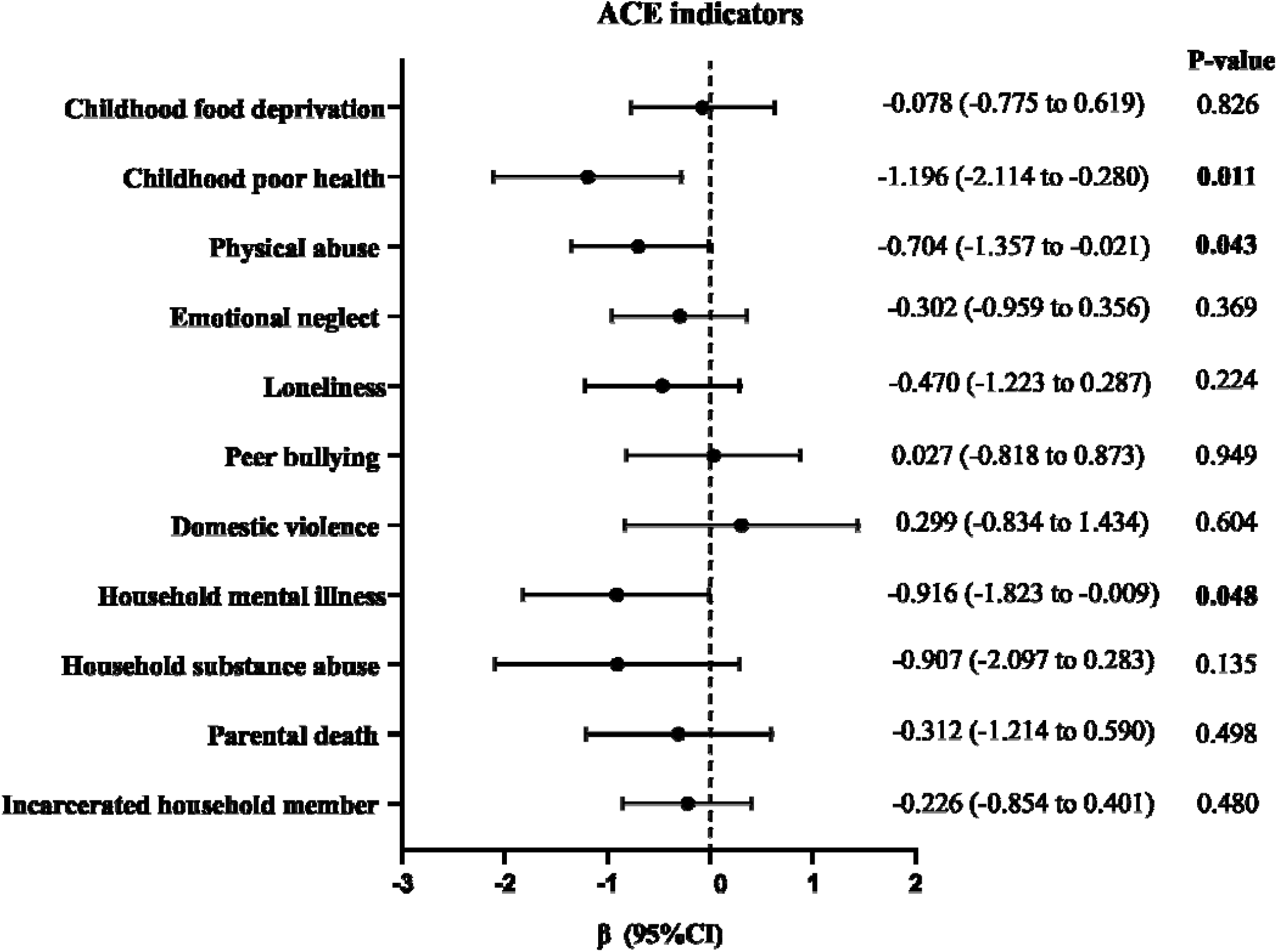
Association between each ACE indicator exposure with reduced kidney function later in life^#^. # Model adjusted for gender, age, BMI, educational level, marriage status, financial situation before 17 years,cigarette smoking status, and alcohol use, hypertension, diabetes and heart disease.

**Table 2.** Association of accumulated ACEs exposure with renal function in later life.

| eGFR | ACE score |  |  |  | P for trend |
| --- | --- | --- | --- | --- | --- |
|  | ≤ 1 | 2 | 3 | ≥ 4 |  |
| Model □ | Ref | -1.798<br>(-2.751 to -0.846)** | -1.351<br>(-2.395 to -0.307)* | -2.548<br>(-3.698 to -1.398)*** | 0.001 |
| Model □ | Ref | -0.404<br>(-1.183 to 0.375) | -0.450<br>(-1.307 to 0.407) | -1.225<br>(-2.170 to -0.280)* | 0.017 |
| Model □ | Ref | -0.379<br>(-1.158 to 0.399) | -0.430<br>(-1.286 to 0.426) | -1.169<br>(-2.113 to -0.225)* | 0.023 |
| CKD |  |  |  |  |  |
| Model □ | Ref | 1.08<br>(0.88 to 1.34) | 1.00<br>(0.80 to 1.27) | 1.38<br>(1.09 to 1.74)** | 0.033 |
| Model □ | Ref | 1.01<br>(0.80 to 1.27) | 1.01<br>(0.78 to 1.30) | 1.38<br>(1.07 to 1.79)* | 0.037 |
| Model □ | Ref | 1.01<br>(0.81 to 1.27) | 1.00<br>(0.77 to 1.29) | 1.35<br>(1.04 to 1.75)* | 0.048 |

Note: eGFR, estimated glomerular filtration rate; CKD, chronic kidney disease.
Model □: crude model
Model □: adjusted for gender, age, BMI, educational level, marriage status, childhood financial status before 17 years, cigarette smoking status, and alcohol use.
Model □: further adjusted for hypertension, diabetes and heart disease based on Model □.

Stratified analysis showed that the association was more pronounced in men, and those who aged ≥ 60 years. The adjusted β and 95%CIs for those who experienced four or more ACEs were -1.561(−2.988 to -0.132) in men and -1.782 (−3.081 to -0.484) for those aged ≥ 60 years, compared with the reference group (Table 3). When stratification by educational level or childhood financial status before 17 years, we observed a higher risk of reduced kidney function in those with lower education, of worse family’s financial status during childhood (Table 3). Trend analysis also showed a significant dose-response relationship. However, we did not find a statistical significance between two groups when stratified by these sociodemographic variables in each ACE group (Table 3).

**Table 3.** Associations of accumulated ACE exposure with reduced kidney function stratified by several sociodemographic variables.

| Variables | ACEs score |  |  |  | P for trend |
| --- | --- | --- | --- | --- | --- |
|  | ≤ 1 | 2 | 3 | ≥ 4 |  |
| Gender |  |  |  |  |  |
| Men | Ref | -0.134<br>(-1.369 to 1.101) | -0.715<br>(-2.036 to 0.607) | -1.561<br>(-2.988 to -0.132)<sup>*</sup> | **0.026** |
| Women | Ref | -0.544<br>(-1.532 to 0.443) | -0.129<br>(-1.242 to 0.983) | -0.766<br>(-2.021 to 0.488) | 0.351 |
| P-value<sup>a</sup> | — | 0.624 | 0.517 | 0.419 | — |
| Age |  |  |  |  |  |
| ≥ 60 years | Ref | -0.161<br>(-1.209 to 0.887) | -0.369<br>(-1.524 to 0.785) | -1.782<br>(-3.081 to -0.484)<sup>**</sup> | **0.017** |
| < 60 years | Ref | -0.628<br>(-1.757 to 0.500) | -0.618<br>(-1.860 to 0.623) | -0.786<br>(-2.137 to 0.565) | 0.248 |
| P-value<sup>a</sup> | — | 0.564 | 0.786 | 0.301 | — |
| Educational level |  |  |  |  |  |
| ≥ Middle school | Ref | 0.132<br>(-1.218 to 1.481) | -0.696<br>(-2.165 to 0.772) | -0.789<br>(-2.459 to 0.880) | 0.240 |
| < Middle school | Ref | -0.633<br>(-1.589 to 0.322) | -0.399<br>(-1.455 to 0.657) | -1.349<br>(-2.498 to -0.201)<sup>*</sup> | **0.045** |
| P-value<sup>a</sup> | — | 0.371 | 0.760 | 0.600 | — |
| Financial situation before 17 years |  |  |  |  |  |
| Same as or better than others | Ref | -0.049<br>(-1.023 to 0.926) | 0.101<br>(-1.007 to 1.209) | -0.796<br>(-2.071 to 0.478) | 0.390 |
| Worse than others | Ref | -1.045<br>(-2.348 to 0.257) | -1.314<br>(-2.680 to 0.053) | -1.855<br>(-3.293 to -0.417)<sup>*</sup> | **0.011** |
| P-value<sup>a</sup> | — | 0.232 | 0.115 | 0.284 | — |

\* $p < 0.05$ , \*\* $p < 0.01$
a: indicated the statistical testing between groups. The bold value indicated statistical significant.

## Discussion

In this national population-based study, the results suggested that early-life exposure to higher ACEs were significantly related to the increased risk of reduced kidney function later in life. The specific individual ACE indicator, especially childhood poor health, physical abuse, and household mental illness might play an important role on the risk of kidney disease across the life course. A significant dose-response relationship between the accumulated ACEs and subsequent CKD was observed. Our findings showed that men who aged ≥ 60 years, with lower educational level, and poorer childhood financial status might have a higher risk of subsequent CKD.

For the significant association between accumulated ACEs exposure with subsequent kidney disease, the results supported the previous literature on how early trauma or childhood adversity could influence the health later in life(13). Consistent with a previous study(14), our study indicated that those who experienced higher ACEs increased the risk of CKD and reduced eGFR in later life. Another study(9)conducted in North Carolina also showed that individuals experiencing four or more ACEs had a higher risk of kidney disease compared to those with 1 ACE, despite the cultural difference. Besides the conventional ACE indicators, we also evaluated several extended indicators including childhood poor health and childhood food deprivation in the current study. Our findings indicated that individuals with poor health during childhood had a higher risk of subsequent kidney disease, similar to previous studies(23, 24). Furthermore, our results showed that experiencing physical abuse or having household mental illness might enhance the risk of kidney disease in later life, which in line with a previous study(25). Since ACEs do not occur in isolation, but rather tend to co-occur among children(9). Individuals who living with parents who have a mental illness tend to receive inadequate physical or emotional care and easily be influenced by substance abuse from their parents, all of which can increase the risk of chronic diseases later in life.

To date, the exact mechanisms are not clear. One possible explanation was that prolonged stress caused by ACEs might induce chronic activation of the hypothalamic -pituitary-adrenal (HPA) axis and then increased allostatic load and affected the regulation of endocrine, immune and autonomic nervous(9). Another likely explanation was that adversity in early life could manifest in alterations in immune system, the expression of certain genes, and increased chronic inflammatory, which associated with various chronic health conditions across the life time (13, 28, 29).

In our study, we found that men with higher ACEs score were more likely to have a higher risk of reduced kidney function in later life, compared with women. One reason behind might be that young boys were often more naughty than girls, and they exposed to a higher number of ACEs in early-life. Another explanation from animal studies revealed that women fetuses might be less vulnerable in adverse utero environments(30). It was reported that placenta of women fetuses had higher 11β-hydroxysteroid dehydrogenase type 2 (11-HSD2) level and lower level of maternal glucocorticoids than men, which could be a underlying reason for reduced nephropathy in male adults (31). Additionally, we observed that individuals who aged 60 years or older having a higher risk of CKD. This was similar to previous publication(32), which found the kidney function decline and CKD were more common in the elderly population. Moreover, We observed significant relationship of the risk of subsequent kidney disease in those with lower education level or worse financial status during childhood when exposure to ACEs. The potential reason behind was that higher education or financial status might mitigate the harmful health effects of ACEs exposure(32). Future studies are needed to confirm our findings.

### Strengths and limitations

Our study has several strengths. First, this was one of the few studies in a developing country to explore a comprehensive range of ACE indicators exposure and subsequent kidney disease by a nationally representative sample, which provided important implications that critical and persisting influence of childhood advertises might last for the life course. Second, the findings indicated that certain predictive factors such as childhood poor health,physical abuse, and household mental illness, which might help policy-makers or researchers to make intervention strategies to address kidney health later in life. Third, we used the serum CysC concentration from laboratory testing to estimate the eGFR to help obtain a more accurate assessment, compared with previous studies which based on self-reported outcomes(9, 14). Last, we evaluated the modification of the association by several sociodemographic variables, which helped identify the vulnerable characteristics.

Despite the novelty of the study, several limitations should be acknowledged. First, the measurements of ACE indicators were based on self-report in the 2014 life history survey, which could lead to recall bias. Second, limited by the CHARLS project, the study population were middle-aged or older Chinese adults, which might induce possible limitation with respect to generalizability of the findings to a more general population. Third, some other ACE indicators, such as sexual abuse and parental separation or divorce were not evaluated due to the unavailable data, despite a wide range of indicators being assessed in this study. Fourth, given the lack of information about the frequency, intensity, or chronicity of ACEs, restricted our ability to analyze the association. Last, the information about potential confounders including dietary factors, air pollution and medical care was not available, which might influence the results. For all these limitations mentioned above, future cohort studies with a large sample size are warranted to validate our findings.

## Conclusions

Our findings suggest that accumulated ACEs exposure in early life significantly associated with an increased risk of kidney disease among middle-aged or older Chinese adults. The association is more pronounced in men, those who aged ≥ 60 years. Furthermore, those with lower educational level or having poor family’s financial status may enhance the risk of decreased kidney function when exposure to ACEs. Considering China’s rapidly aging population and high prevalence of CKD, our study provides important public health implications for reducing childhood adversities and promoting kidney health in later life.

## Data Availability

All data produced in the present study are available upon reasonable request to the authors.

